# Changes to an intensive care unit *Acinetobacter baumannii* population following COVID-19 disruptions and targeted infection prevention interventions

**DOI:** 10.1101/2024.02.02.24302174

**Authors:** Haiyang Liu, Robert A. Moran, Emma L. Doughty, Xiaoting Hua, Ann E. Snaith, Linghong Zhang, Xiangping Chen, Feng Guo, Willem van Schaik, Alan McNally, Yunsong Yu

## Abstract

Carbapenem-resistant *Acinetobacter baumannii* (CRAB) is a persistent nosocomial pathogen that poses a significant threat to global public health. A three-month cross-sectional observational study was conducted in a 28-bed ICU in Hangzhou, China. The same ICU was sampled for the same duration and with a similar methodology in 2019, 20 months prior to the outset of this study. Following COVID-19-associated delays, a series of IPC measures targeting patients, staff and the ICU environment were implemented for 8 months prior to and throughout this study. A total of 5,341 samples were collected from the ICU environment (n = 4450) and patients (n = 891). *A. baumannii* isolates were obtained from 9·5% of these samples (n = 505). Most *A. baumannii* isolated in this ICU were CRAB (419/518; 80·9%). Fewer CRAB were isolated here (407 from 363 sampling occasions) than in 2019 (502 from 336 sampling occasions). However, MIC_50_/MIC_90_ values for imipenem increased from 32/64 mg/L in the 2019 study to 64/128 mg/L here. This was accompanied by the proportion of global clone 2 (GC2) isolates falling from 99·5% in 2019 to 50·8% (213/419) in 2021. The phylogenetic diversity of GC2 increased, apparently driven by regular introductions of distinct clusters in association with patients. The remaining CRAB (40·2%; 206/419) were a highly clonal population of ST164, which appears to have persisted in the ICU since an introduction in mid-2020. We found clusters of GC2 and ST164 isolates with identical core genomes in different room or bed unit environments, and in multiple patients, indicative of transmission in the ICU.

Changes to IPC procedures in this ICU were associated with a reduction in the total prevalence of CRAB, and in the number of CRAB isolated from clinical specimens. At the phenotypic level, the CRAB population exhibited increased resistance to carbapenems, and this may be the result of increased antibiotic prescribing over the COVID-19 period. The increased diversity of this CRAB population appears to have been the result of repeated introductions to the ICU with patients, which have continued despite interventions.

## Introduction

*Acinetobacter baumannii* is a Gram-negative bacterial pathogen that causes ventilator-associated pneumonia and bloodstream infections in critically ill hospital patients globally.^1,2^ The morbidity and mortality associated with *A. baumannii* infections are exacerbated by its extensive resistance to antibiotics. Carbapenem-resistant *A. baumannii* (CRAB) are often only sensitive to tigecycline and polymyxins,^3^ and can persist in hospital environments and patients for extended periods.^1^ Resolving CRAB outbreaks can require modifications to practice and infrastructure that impose significant clinical, logistical, and financial costs.^4^ In 2018, the World Health Organisation declared CRAB a priority organism for which novel therapeutics are urgently required.^5^

Given the limited options for antimicrobial chemotherapy, infection prevention and control (IPC) strategies must play a substantial role in limiting the impact of CRAB in hospitals.^6^ Effective genomic surveillance has the capacity to inform IPC approaches by defining outbreaks and identifying sources and pathways for bacterial transmission. Recent hospital studies have demonstrated the utility of whole-genome sequencing (WGS) for high-resolution characterisation of *A. baumannii* outbreaks or persistent populations.^6–9^ The crucial need to develop a thorough understanding of the longer-term persistence, transmission and evolution of CRAB populations in nosocomial environments provided the motivation for undertaking this study.

In August-October 2019, we conducted a 13-week genomic surveillance study of CRAB in a 28-bed intensive care unit (ICU) located in Hangzhou, China.^10^ We found that the ICU CRAB population was dominated by representatives of global clone 2 (GC2), which is one of two clones that account for the majority of CRAB globally.^3^ Consistent with global reports, carbapenem resistance in this GC2 population was conferred by the *bla*_OXA-23_ carbapenemase gene, which was present in composite transposons Tn*2006* or Tn*2009* inserted at several different chromosomal sites.^10^ Comparative analyses revealed that the GC2 population in this ICU was phylogenetically diverse, and the diversity appeared to have been shaped by independent introductions of multiple distinct GC2 clusters to the ICU. We detected putative CRAB transmission events between bed units and rooms within the ICU, and the spatiotemporal co-location of diverse CRAB clusters facilitated three horizontal gene transfer events over the course of the study.^10^

Informed by that first study, we planned to implement a set of IPC interventions, then perform a follow-up study to evaluate their effectiveness in reducing the prevalence of CRAB in the ICU. However, ICU operations were interrupted by the outbreak of COVID-19 in early 2020, and access to the ICU was restricted. Consequently, the implementation of planned CRAB-focused interventions was delayed until September 2020. Once introduced, the suite of interventions targeted ICU patients, the ICU environment (including equipment and sinks), and ICU staff (Table S1). Full access to the ICU was restored in late November 2020, enabling us to conduct a second high-resolution genomic surveillance study, with a methodology identical to that of our pre-COVID-19 pandemic study, in May-July 2021.

Here we describe the follow-up cross-sectional observational study of *A. baumannii* in this Hangzhou ICU. Over a 13-week period, we applied a deep-sampling approach to ICU patients and the ICU environment. All *A. baumannii* isolates were whole-genome sequenced. The structure and dynamics of this population were compared to those from our original study to evaluate the population’s longer-term development, and its response to IPC interventions.

## Methods

### Consent and research ethics

Ethical approval and informed consent were obtained by the Sir Run Run Shaw Hospital (SRRSH) Zhejiang University local ethics committee (approval number 20190802-1). This work was part of a study registered as a clinical trial with ClinicalTrials.gov (NCT04310722).

### IPC interventions implemented before this study

A bundled set of interventions that aimed to address the abundance of CRAB in this ICU were implemented 8 months prior to the outset of this study. The interventions included changes to daily practice in the ICU, alongside raising staff awareness around the issue. The interventions are listed in Table S1. Briefly, patient care was modified by introducing “water-free” cleaning methods and by installing disinfection devices for liquid waste. ICU sinks ceased operation. Environmental cleaning was performed more regularly, with particular attention paid to end-of-stay cleaning procedures and communal surfaces such as keyboards used routinely by ICU staff. Education for staff was enhanced, and enforcement of hand hygiene practices was targeted through compliance monitoring and the introduction of a compliance management system.

### Study design and sample collection

This cross-sectional observational study was conducted in a 28-bed ICU in SRRSH, Hangzhou, China from May to July 2021. A map of the ICU is presented in Figure S1. Unless sampling was perceived to be medically detrimental, ICU patient sampling was planned for the beginning of the study or on admission to the ward and weekly thereafter. Patient samples were routinely collected from oral and rectal swabs, and from nasogastric, nasojejunal, endotracheal, or tracheostomy tube swabs when present. Clinical samples were obtained from patients as part of normal medical practice, and all *A. baumannii* obtained from clinical samples over the study period were retained. Equipment, sinks and other surfaces within patient rooms were swabbed weekly. Equipment, surfaces, and sinks in communal areas outside patient rooms were sampled monthly.

### Sample processing and DNA sequencing

Environmental surfaces (approximately 10 x 10 cm) were swabbed with sterile swabs (COPAN, Italy) pre-moistened with 2 mL TSB supplemented with 0·1% sodium thiosulfate (Sangon Biotech, China). The swabs were immediately placed into 14-mL sterile tubes containing 2 mL TSB with 0·1% sodium thiosulfate then incubated for 24 hours at 37°C. After culture, 20 µL overnight bacteria were streaked onto Acinetobacter CHROMagar plates (CHROMagar, Paris, France) followed by overnight incubation at 37°C.

For patient samples, rectal swabs containing Cary-Blair Transport Medium (Gongdong, Taizhou, China) were used and other swabs were moistened using 0·9% saline before sampling. These swabs were plated directly onto Acinetobacter CHROMagar plates and incubated at 37°C for 24 hours. Based on red colour and morphology, a single, isolated colony of presumptive *A. baumannii* was selected and streaked onto a Mueller-Hinton (MH) agar plate (Oxoid, Hampshire, UK), then incubated again at 37°C for 24 hours. A single, isolated colony from the MH plate was selected and stored for downstream analyses.

Species identification was confirmed using Matrix-assisted laser desorption ionization-time of flight mass spectrometry (MALDI-TOF MS) (bioMérieux, France) and 16S rRNA gene sequencing using the primers 27F-5’-AGAGTTTGATCCTGGCTCAG-3’ and 1492R-5’-GGTTACCTTGTTACGACTT-3’.

### Antimicrobial susceptibility profiling

Minimum inhibitory concentrations (MICs) of meropenem, imipenem, polymixin B, tigecycline, cefoperazone/sulbactam (1:1), cefoperazone/sulbactam (2:1), sulbactam, amikacin and ciprofloxacin were measured using the broth microdilution method of plate for “Susceptibility testing of non-fastidious Gram Negative isolates”. Results were interpreted according to the Clinical and Laboratory Standards Institute 2021 guidelines and European Committee on Antimicrobial Susceptibility Testing (EUCAST) 2023 breakpoint tables (http://www.eucast.org/clinical_breakpoints/) for polymyxin B (S: ≤2 mg/L, R: >2 mg/L). Tigecycline results were interpreted based on Food and Drug Administration (FDA) guidelines. *Escherichia coli* ATCC 25922 was used as the quality control strain.

### Illumina sequencing

Collected isolates were subjected to Illumina sequencing. One colony of each isolate was cultured in 2 mL Mueller-Hinton broth (Oxoid, UK) for 24 hours at 37°C. Cell pellets were harvested and DNA was isolated using the Qiagen Mini Kit (Qiagen, Germany). The quality and quantity of DNA were assessed using a NanoDrop 2000 (Thermo Scientific, USA) and Qubit 4.0 fluorometer (Invitrogen, USA).

Libraries were prepared using the TruePrepTM DNA Library Prep Kit V2 (Vazyme) according to the manufacturer’s protocol. Individual libraries were assessed on the QIAxcel Advanced Automatic nucleic acid analyzer using a high-resolution gel cartridge (Qiagen, Germany), and then were quantified by qPCR using the use of KAPA SYBR FAST qPCR Kits Kapa KK4610 (KAPA Biosystem, Wilmington, MA, U.S.A.). Paired-end sequencing (2×150-bp reads) was performed on the Illumina HiSeq X Ten platform (Illumina Inc., San Diego, CA, USA).

### Oxford Nanopore sequencing

Long-read sequencing was performed for 60 isolates chosen after Illumina sequencing on the basis of phylogenetic, resistance gene and plasmid diversity. Genomic DNA was extracted using the Gentra® Puregene® Yeast/Bact. Kit (Qiagen, Germany) according to the manufacturer’s protocol with minor modifications. Briefly, cell pellets were harvested in a sterile 15 mL falcon tube from 5mL overnight culture. Each pellet was resuspended in 500 µL Lysis Buffer Solution, transferred to a Phase Lock tube and incubated at 37°C for 30 minutes. Cell lysates were treated with 2 µL RNase A (100 mg/mL, Qiagen, Germany), gently mixed by flicking the tube and incubated at 37°C for 30 minutes. The cell lysate was further treated with 3 µL Proteinase K (Qiagen, Germany), mixed by gentle inversion and incubated at 50°C for 90 minutes. The suspension was mixed with 500 µL phenol:chloroform:isoamyl alcohol (25:24:1) by inversion and centrifuged at 20,000 rcf for 10 minutes. The supernatant was collected and DNA was purified using AMPure XP beads (Beckman Coulter™) with minor modifications to the manufacturer’s protocol: 70% ethanol was used for washing the DNA on the beads before incubation at 37°C for 5 min to dry off residual ethanol. The DNA was resuspended in 150 µL of nuclease-free water. The quality and quantity of DNA were assessed as described above.

Oxford Nanopore sequencing libraries were prepared using the SQU-LSK109 Ligation Sequencing Kit (Oxford Nanopore Technologies, UK) in conjunction with the PCR-Free ONT EXP-NBD104 Native Barcode Expansion Kit (Oxford Nanopore Technologies, UK) according to the native barcoding genomic DNA protocol. DNA was processed without the optional shearing steps to select for long reads. After quantification of the individual libraries by Qubit and normalisation of library concentrations, the library was sequenced on the GridION X5 platform (Oxford Nanopore Technologies, UK).

Raw Illumina and Minion sequence data for all isolates is available via National Center for Biotechnology Information under BioProject accession PRJNA1034534.

### Bioinformatic analyses

Sequence reads were trimmed, assembled and assessed for quality. MLST (https://github.com/tseemann/mlst) was used to determine multi-locus sequence types with the Pasteur and Oxford typing schemes.^11,12^ Typing of capsular polysaccharide (KL) and lipooligosaccharide outer core (OCL) synthesis loci was conducted with Kaptive.^13^ AMRFinder was used to identify antimicrobial resistance genes.^14^ Plasmid replicons were typed using the Hamidian lab typing scheme.^15^

Snippy v4.4.5 (https://github.com/tseemann/snippy) was used to align Illumina reads against a corresponding reference hybrid assembly and generate a core genome alignment. Polymorphic sites were extracted with Gubbins v2.4.0 excluding those that were predicted to occur via recombination.^16^ Phylogenies were constructed from these polymorphic sites using IQtree v2.0.3. Divergence dating was undertaken with the least-squares method implemented by IQTree v2.0.3, using the previously generated tree, Gubbins fasta file, and a GTR+G model.^17,18^ SNP-distances were calculated from the Gubbins-filtered polymorphic sites file using SNP-dists 0.6.3 (https://github.com/tseemann/snp-dists).

## Results

### *A. baumannii* infections were rare despite high abundance in patients and bed units

A total of 131 patients (79 male; 52 female; median age 68 years; interquartile range [IQR] = 55·5 – 77·5) were sampled over the three-month study period (Table S2). The median length of patient stay, from admission to discharge or the end of the study, was 10 days (IQR: 5 – 21·5 days). Most patients (53·4%) were in the ICU for ≤10 days, but 17·1% were present for >31 days. Samples were taken each Tuesday, and patients were screened within the first 48 hours after admission when possible. “Bed units”, defined as the environmental sites of each bed and its associated equipment, were sampled on 363/364 (99·7%) planned sampling occasions, as one patient was undergoing bedside surgery on a single planned sampling occasion. Patients were sampled on 303/346 (87·6%) planned occasions.

A total of 5,341 samples were collected from patients and environmental sites. *A. baumannii* was isolated from 505 samples: 382/4450 (8·6%) environmental samples and 123/891 (13·8%) patient samples. Bed units yielded more environmental isolates (374/4317, 8·7%) than communal areas outside bed units (8/133 samples, 6·0 %). Within bed units, ventilators (56/281 samples, 19·9%) and ventilator shelves (40/275 samples, 14·6%) yielded the most *A. baumannii*. Of the 131 patients screened, 41·2% (54/131) were *A. baumannii*-positive on at least one sampling occasion, with most of the positive samples originating from nasogastric tube (33/174, 19·0%) or oral (40/220, 18.2%) swabs. A set of 13 isolates obtained from diagnostic clinical specimens were collected from 10 patients over the study period; one from an abdominal excision exudate and 12 from sputum.

### Most ICU *A. baumannii* were carbapenem-resistant

Over the study period, 19·1% (25/131) of ICU patients were treated with the carbapenems imipenem, meropenem, or ertapenem. MIC testing of all 520 *A. baumannii* in this collection revealed that the rates of resistance to both imipenem and meropenem were 80·9% (419/518) (Figure S2). A larger proportion of patient isolates (116/123; 94.3%) were CRAB than environmental isolates (303/382; 79.3%). The β-lactam/β-lactamase inhibitor combinations cefoperazone/sulbactam and piperacillin/tazobactam were administered to 14·5% (19/131) and 40·5% (53/131) of patients, respectively. The resistance rates to these combinations were 80·3% (416/518) for cefoperazone-sulbactam (1:1) and 81·1% (420/518) for cefoperazone-sulbactam (2:1). Resistance rates to sulbactam alone, ciprofloxacin, and amikacin were 80·9% (419/518), 80·9% (419/518), and 23·0% (119/518), respectively. All isolates were sensitive to colistin and to tigecycline.

To quantify levels of carbapenem resistance in the CRAB populations from before and after the intervention period, we compared the imipenem MICs that would be sufficient to inhibit 50% (MIC_50_) and 90% (MIC_90_) of all isolates in each collection. For the 419 CRAB isolated in this study, the MIC_50_ and MIC_90_ to imipenem were 64 mg/L and 128 mg/L, respectively. The imipenem MIC_50_/MIC_90_ values for the 551 CRAB isolated in our first study were 32 mg/L and 64 mg/L, respectively. This data reflects an increase in carbapenem resistance levels in this ICU between 2019 and 2021. However, total sensitivity to colistin and tigecycline remained consistent across study periods.

### Whole-genome sequencing revealed shifts in the circulating CRAB population

All *A. baumannii* isolates were whole-genome sequenced (Table S3). Genome sequence data revealed that the population included representatives of 35 different sequence types according to the Pasteur MLST scheme (Figure 1A). Most of the diversity was seen amongst the carbapenem-sensitive population, which included isolates from 33 different STs, 20 of which were represented by a single isolate each. Only three of these STs were represented by ten or more isolates: ST240, ST40, and ST221. The carbapenem-resistant population was comprised of just two sequence types: ST2/GC2 (213 isolates; 50·8%) and ST164 (206 isolates; 49·2%). The presence of ST164, accounting for approximately half of the CRAB isolates in this study, represents a major shift from 2019, where the CRAB population was dominated by GC2 (548/551; 99·5%) (Figure 1A). ST164 isolates were obtained from clinical specimens in this ICU earlier in 2021,^19^ and this ST has recently been declared an international clone (IC11), with reports of human infections in Europe, Asia, Africa and the Middle East.^20^ Imipenem MIC_50_/MIC_90_ values for the 206 ST164 isolates were 128/128 mg/L, and 32/64 mg/L for the 213 GC isolates, indicating that the appearance of ST164 was responsible for the overall increase in carbapenem resistance between studies.

**Figure 1:**
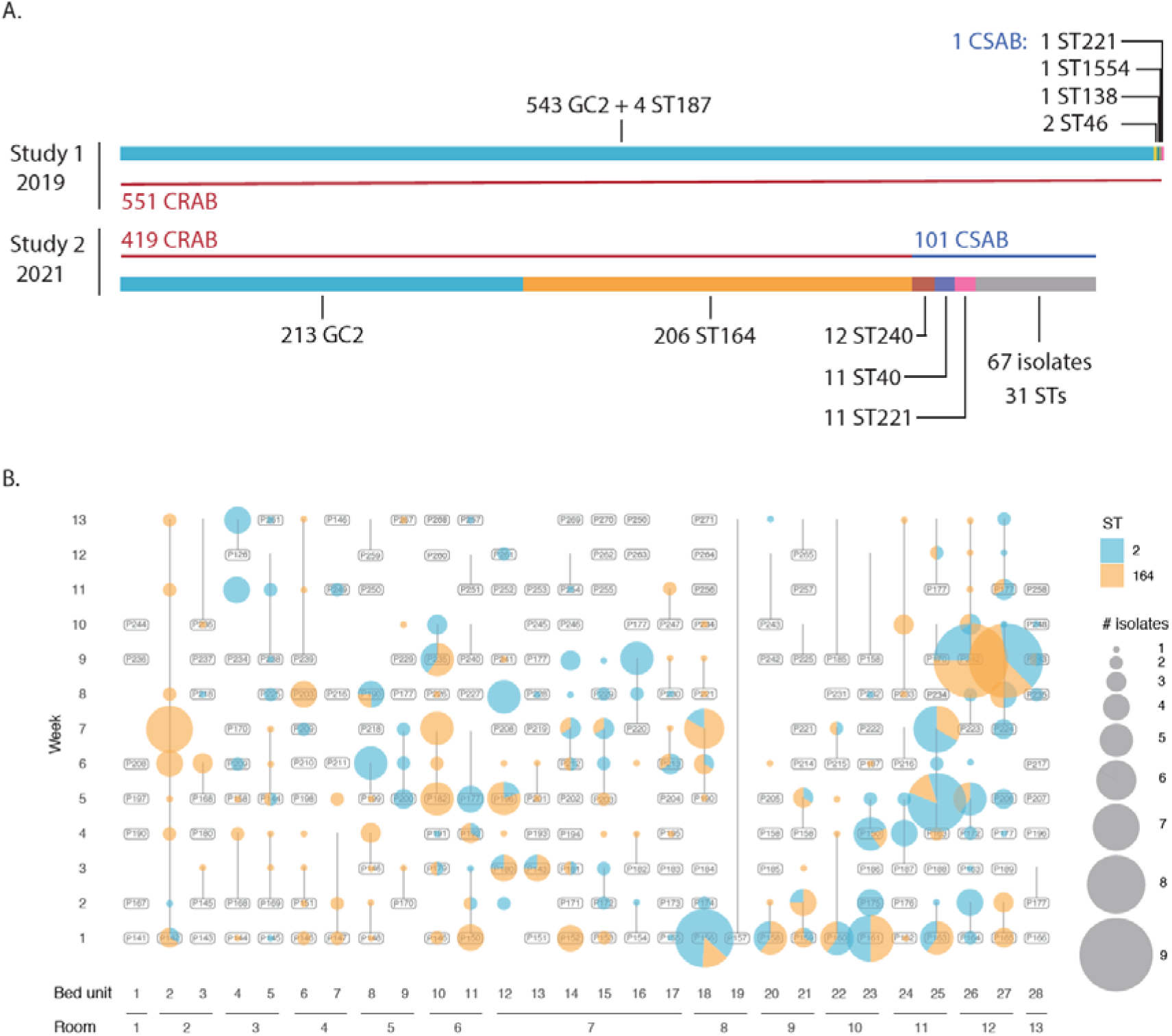
The ICU *A. baumannii* population. A) Comparison of the *A. baumannii* populations collected in this ICU over the initial (2019) and follow-up (2021) study periods. Horizontal bars are proportional to the number of isolates of given sequence types (STs), which are labelled and represented by different colours. Thinner horizontal lines and labels indicate whether isolates were carbapenem-resistant (CRAB; red) or carbapenem-sensitive (CSAB; blue). B) Overview of the spatiotemporal distribution of GC2/ST2 (blue) and ST164 (orange) isolates over the course of the 2021 study. Bed unit and room numbers are indicated on the horizontal axis, and study week numbers on the vertical axis. The sizes of circles correspond to the number of isolates collected from each patient/bed unit at each sampling timepoint.

GC2 and ST164 were isolated throughout the study period and across the ICU (Figure 1). On 37/364 (10·2%) sampling occasions, GC2 and ST164 were isolated from the same bed unit. Despite their near-even prevalence in the ICU environment and patients, GC2 strikingly accounted for 12/13 (92·3%) clinical isolates obtained over the study period, with the remaining sputum isolate belonging to ST164. In contrast to GC2 and ST164, carbapenem-sensitive (CSAB) STs appeared sporadically over the course of the study and were generally localised to a single room, patient or time point (Figure S3).

### The GC2 population shifted significantly between studies through the introduction of multiple discrete sub-clusters

Consistent with our first study, carbapenem-resistance in GC2 CRAB isolates was conferred by the carbapenemase gene *bla*_OXA-23_. To determine the genomic contexts of these *bla*_OXA-23_ genes, we examined 19 hybrid-assembled complete GC2 genomes, and screened the collection of 213 GC2 draft genomes with signature sequences indicative of the Tn*2006* or Tn*2009* insertions identified in our first study.^10^ This revealed that the chromosomal positions encountered in the first study accounted for *bla*_OXA-23_ carriage in all GC2 isolates collected here (Figure 2A; Table S3). Additionally, one hybrid-assembled genome (DETAB-E2110) contained two copies of Tn*2006*, with the second copy found in the 13,545 bp plasmid pDETAB10::Tn*2006* (Figure S4). pDETAB10::Tn*2006* was generated by the insertion of Tn*2006* into the backbone of the 8,731 bp R2-T1 type plasmid pDETAB10, which was present in 75/547 (13·7%) GC2 isolates in our first study and 175/213 (82·2%) GC2 isolates here. Screening this collection with the signatures of this insertion revealed that pDETAB10::Tn*2006* was not present in any other isolates. DETAB-E2110 had MIC values of 128 mg/L and >128 mg/L for imipenem and meropenem, which were the equal-highest observed in this collection.

**Figure 2:**
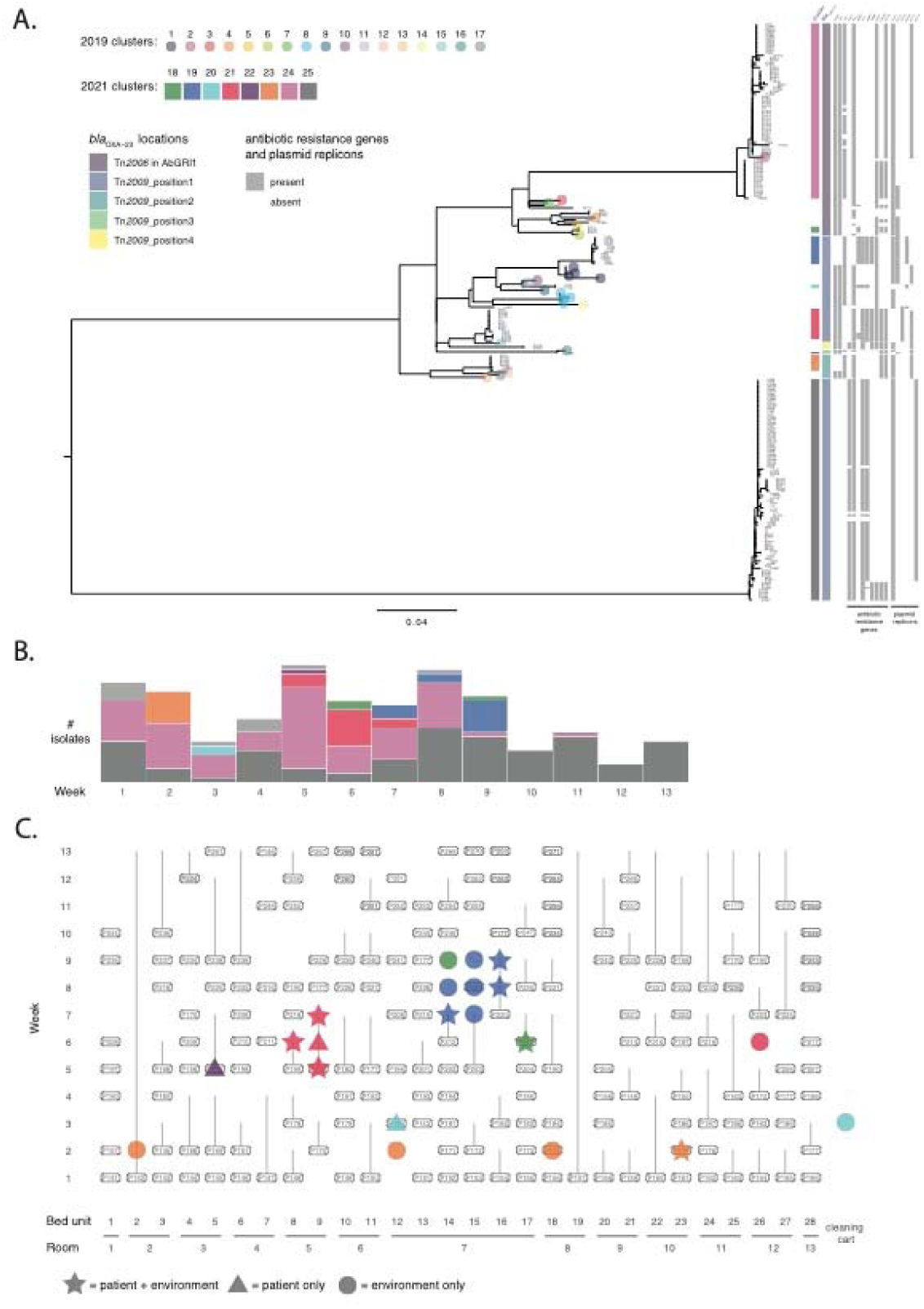
The ICU GC2 population observed in this study. A) Core-gene phylogeny (left) and cluster characteristics (right). Reference genomes for clusters obtained in the 2019 study are indicated by coloured circles at branch tips. Cluster designations from this study are indicated by coloured boxes to the right of the phylogeny, along with boxes shaded as outlined in the key to the left of the figure. B) Temporal distribution of GC2 clusters over the course of this study. For each study week, columns are divided according to the numbers of isolates of each GC2 cluster that were obtained, with colours corresponding to the key in part A. C) Spatiotemporal distribution of transient GC2 clusters introduced by patients over the course of this study. The meaning of shapes is outlined in the key below, and colours correspond to those in the key in part A.

Although the GC2 isolates collected here shared Tn*2006*/Tn*2009* insertions with the GC2 clusters examined in our previous study, chromosomal segments that contain transposons can be exchanged by homologous recombination between distinct *A. baumannii* strains,^21^ so we sought phylogenetic evidence to assess cluster persistence in this ICU. To determine whether the GC2 population observed in this study was derived from that present in 2019, we constructed a phylogeny from the GC2 genomes obtained here, along with representative genomes for each of the 17 GC2 clusters identified in our first study (Figure 2A). The phylogeny featured three large clusters that included 49, 72 and 92 GC2 genomes from this study. Representatives of 16 GC2 clusters from the 2019 study were confined to the diverse central cluster, with a single representative present in one of the two more clonal clusters, cluster 24 (C24) (Figure 2A). Within the diverse central cluster, representatives from study 1 were largely distinct from isolates collected here, with the exception of C16, which differed from isolate DETAB-P494 by just one core-gene SNP (cgSNP). Overall the GC2 population was more diverse in 2021 than it was in 2019, which is consistent with frequent and ongoing introductions of distinct GC2 clusters to the ICU. Further supporting this, some GC2 clusters appeared briefly in the ICU, and were present for no more than three sampling weeks (Figure 2B). These will be referred to as transient GC2 clusters.

To detect putative patient-associated introductions to the ICU over the course of this study, we identified instances where, apart from in week 1, the first isolates of transient GC2 clusters appeared in patient samples. We found six such instances, involving six different clusters (Figure 2C). Within transient clusters, isolates differed by 0-4 cgSNPs. C22 was represented by a single patient isolate, but isolates of the other five clusters were found in two or more different environments each. These included the bed units occupied by the patients that initially carried the clusters, and either bed units within the same rooms (C18, C19, C21), bed units in different rooms (C23), or a communal cleaning cart (C20) (Figure 2C). C19 and C21 were carried by two patients each, and the second patient that each cluster was isolated from most parsimoniously acquired C19/C21 from their contaminated room environments. All six transient GC2 clusters that first appeared in patient samples disappeared from the ICU following the departure of the last patient that carried them (Figure 2C).

The largest GC2 clusters, C24 and C25, were present in most (10/13 for C24) or all (13/13 for C25) study weeks (Figure 2B). The largest cluster, C25, only contained genomes from this study, all of which differed from one another by 0-6 cgSNPs (Table S4). Isolates in C24 differed from one another by 0-11 cgSNPs, and from the study 1 representative C2 genome by 5-10 cgSNPs (Table S4). As representatives of C24 and C25 were already present in the first week of this study (Figure 2B), we did not capture their introductions to the ICU, so cannot determine whether they were introduced in single or multiple events. However, the presence of sub-clusters of C24 and C25, containing isolates that do not differ at the cgSNP level (0 cgSNPs) but were found in multiple bed units or rooms, is evidence for the ongoing transmission of C24 and C25 within the ICU.

### ST164: A dominant new strain type recently introduced but fully established in the ICU

ST164 was not detected in this ICU in our first study. It was therefore important to establish whether this lineage had been introduced recently or had persisted in the ICU for an extended period. To assess their structural diversity, eight ST164 isolates were hybrid-sequenced to generate complete genomes. All eight complete genomes consisted of a 3.9 Mbp chromosome and five plasmids, pDETEC17 to pDETEC21, that ranged from 12,790 bp to 2,309 bp (Figure 3A). Screening the entire collection of 206 ST164 short-read assemblies revealed that the five plasmids were present in almost all ST164 isolates (pDETAB19 195/206; pDETAB17 201/206; others 206/206).

**Figure 3:**
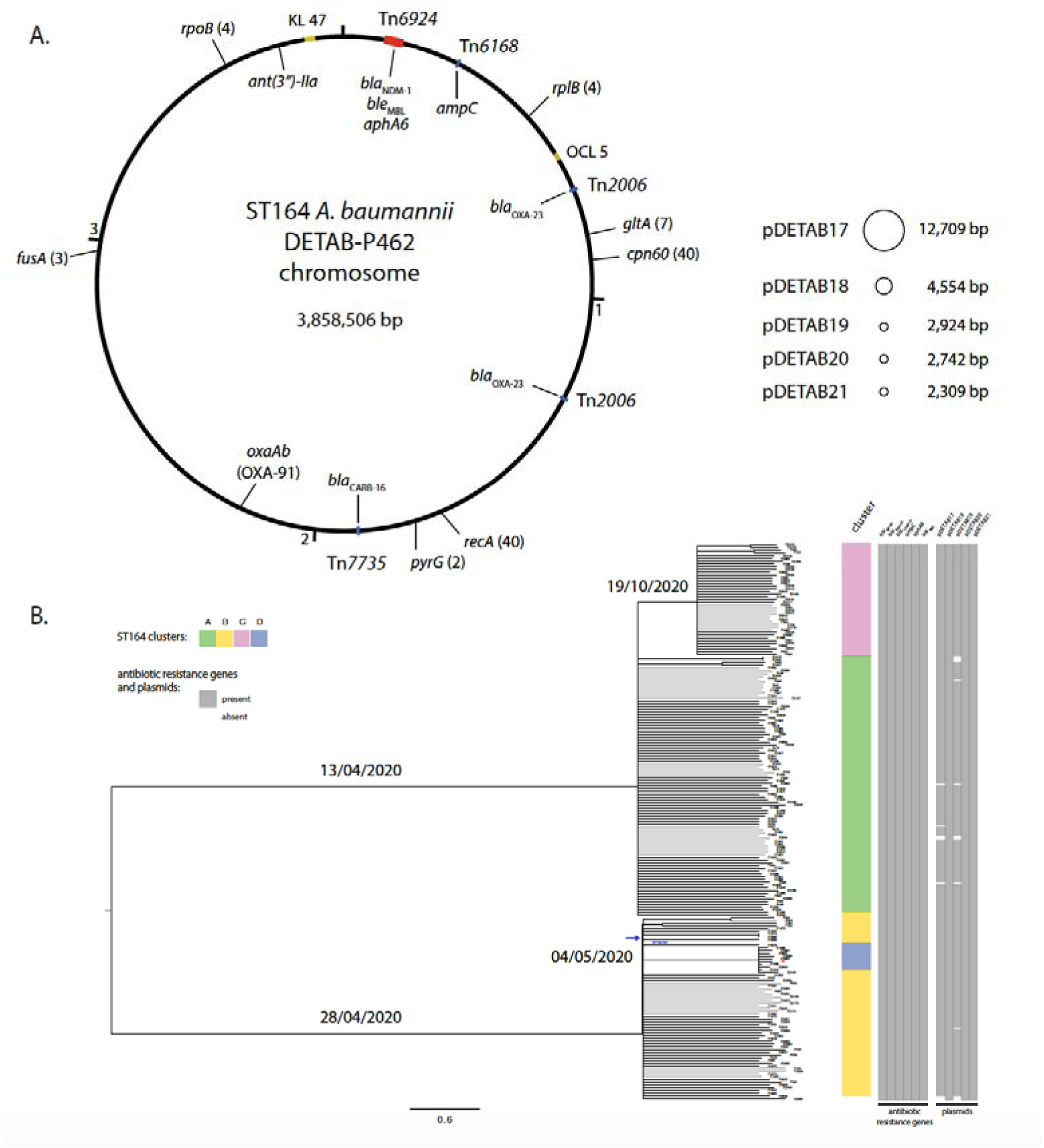
Genomic characteristics of ST164 *A. baumannii*. A) Schematic overview of the genome of ST164 isolate DETAB-P462. The chromosome is shown as a large circle with the locations of antibiotic resistance genes, MLST alleles, capsule (K) and outer core (OC) loci indicated. To the right, plasmids pDETAB17-21 and their sizes are shown (not to scale). B) Time-dated phylogeny of the ST164 population examined in this study. Prominent branch dates are labelled, and the colours to the right of the phylogeny reflect ST164 clusters or the presence of antibiotic resistance genes and plasmids as outlined in the key. The blue arrow indicates the position of DETAB-R21, the reference isolated that was isolated in this ICU in early 2021.

All ST164 genomes contained five acquired antibiotic resistance genes (ARGs). Four of these encoded beta-lactamases: *bla*_NDM-1_, *bla*_OXA-23_, *bla*_CARB-16_ and *ampC* (also called *bla*_ADC_). In the complete ST164 genomes, all ARGs were located in the chromosome (Figure 3A). The *bla*_NDM-1_ gene was in the complex transposon Tn*6924*,^22^ along with the bleomycin resistance gene *ble*_MBL_ and the aminoglycoside resistance gene *aphA6*. The remaining beta-lactamase genes were in four IS*Aba1* composite transposons: *bla*_OXA-23_ in two copies of Tn*2006*, *ampC* in Tn*6168*, and *bla*_CARB-16_ in a novel transposon named Tn*7735* (Figure 3A). Screening draft genomes with the junction sequences associated with all five ARG-containing insertions revealed that they were totally conserved across this ST164 population.

The near-total conservation of mobile genetic elements and chromosomal insertion sites suggested that the ST164 population in this ICU was highly clonal. To confirm this, we determined cgSNPs between all isolates in this collection and found that they ranged from 0-21. We did not find evidence for the introduction of ST164 clusters with patients over the course of this study, and we were aware that the same ST had been isolated from a sputum specimen collected in this ICU in January 2021,^19^ four months prior to this study. We therefore used the genome of that clinical isolate, DETAB-R21, as a reference for a time-dated phylogeny for the genomes generated here (Figure 3B). The time-dated phylogeny was consistent with this ST164 population having diversified from an initial introduction to the ICU in mid-2020. The two deepest branches in this phylogeny had estimated dates of divergence from a common ancestor in April 2020, giving rise to clusters 164-A and 164-B (Figure 3B). DETAB-R21 clustered with 164-B. Both 164-A and 164-B were present in the first and last weeks of this study, but were not present in every intervening week, with no 164-A isolates in week 11 and no 164-B in weeks 3 and 12 (Figure 4). Clusters 164-C (present in weeks 5-13) and 164-D (weeks 2, 5-10), emerged from 164-A and 164-B, respectively (Figure 3B), and were not present in the ICU at the outset of the study (Figure 4). Notably, the first isolates of 164-C and 164-D were derived from environmental samples rather than patient samples, consistent with their emergence from an existing ICU population rather than from patient-associated introductions.

**Figure 4:**
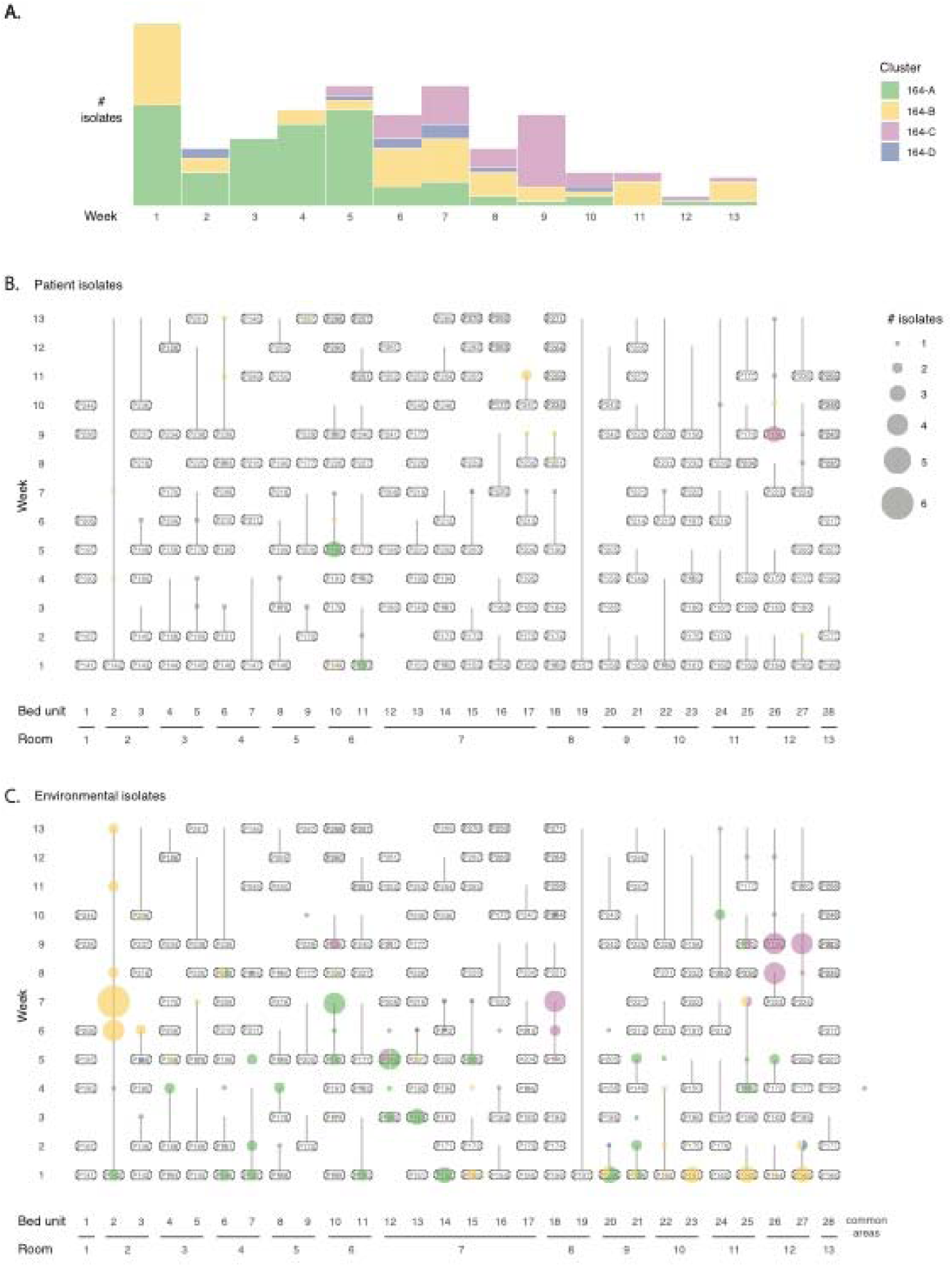
Distribution of ST164 in the ICU. A) Temporal distribution of ST164 clusters over the course of this study. For each study week, columns are divided according to the numbers of isolates of each ST164 cluster that were obtained, with colours corresponding to the key. Spatiotemporal distribution of ST164 clusters isolated from patients (B) and the ICU environment (C). Colours correspond to the key in part A, and the sizes of bubbles to the numbers of isolates indicated by the key in part B.

Over the course of the study, we observed 10 clear instances where patients were admitted to the ICU, were *A. baumannii*-negative at one or more sampling points, but then yielded ST164 from subsequent oral, rectal or tube swabs. In all such cases, patient-derived ST164 isolates were identical (0 cgSNPs; Table S5) to one or more isolates that had previously been obtained from the ICU environment or other ICU patients. We conclude that these events represent acquisition of ST164 from the persistent ICU population. Acquisition events involved three of the four ST164 phylogenetic clusters: ST164-A in 3/10 cases, ST164-B in 4/10, and ST164-C in 3/10. In one further case, a patient that produced GC2 from their first screening samples appeared to acquire ST164-D, which was isolated from a subsequent oral swab and a clinical sputum sample.

### Persistence of CRAB in the ICU results in clinically-relevant CRAB acquisition by patients

Using the phylogenetic evidence described above, we sought to determine the derivation of each of the 13 CRAB clinical isolates collected over the course of this study, and therefore to identify the routes through which they might have been acquired by these 10 patients over the study period. Most (12/13) of the clinical isolates were GC2, and amongst these 10 were C25, one was C24, and one was C21. The remaining clinical isolate was ST164-D. Representatives of all four of these clusters were present in multiple patient and environmental samples prior to their appearance in clinical samples, but to avoid any ambiguity associated with small numbers of cgSNPs (≤4), we focused on comparing identical (0 cgSNP) isolates when describing clinically-relevant CRAB acquisition in the ICU.

The GC2 C21 cluster was clearly introduced to the ICU by patient 200 (P200) (see above, Figure 2), so its presence in a P200 clinical sample is indicative of acquisition elsewhere before the development of clinically-relevant symptoms requiring sputum sample collection in this ICU. The GC2 C24 isolate obtained from P172 was identical to isolates obtained from P172’s first screening samples on arrival to the ICU, which might suggest that they carried C24 on arrival. However, because C24 was circulating in the ICU (see above), and identical isolates had been obtained from other patients and the ICU environment prior to the admission of P172, it is also possible that P172 acquired C24 in the ICU before their first screening samples were taken.

In the remaining eight cases, patients had been CRAB-negative at one or more sampling points prior to the development of symptoms that necessitated clinical sample collection. It therefore seems most parsimonious that all of these patients acquired CRAB in the ICU, and in three of these cases isolates identical to the clinical isolates had previously been isolated from the ICU environment. In the remaining five cases, the clinical isolates were not identical to any other isolates in the collection, but were highly similar to multiple other isolates, differing by ≤4 cgSNPs. We conclude that acquisition in the ICU was responsible for the majority (9 or 10/10) of CRAB found in clinically-relevant patient samples over the course of this study.

## Discussion

This follow-up observational study has allowed us to assess the influence of bundled IPC interventions on an endemic ICU CRAB population, in a region where CRAB prevalence is highest globally.^23^ The nature of bundled intervention strategies complicates assessments of the impacts of individual IPC measures, and this was further complicated here by the time that passed between studies, as well as by the changes to ICU access and practice associated with responses to COVID-19. However, the unique timing of our studies either side of the emergence of COVID-19 also afforded us an opportunity to determine the joint impacts of the intervention strategies as well as the COVID-19 pandemic on antimicrobial resistance in *A. baumannii*.

The early and middle periods of the COVID-19 pandemic were associated with increased rates of antibiotic prescribing in China,^24^ which may have augmented resistance levels in nosocomial bacterial populations. Increased prescription of ß-lactam antibiotics (including carbapenems) and tigecycline, which have been reported across multiple studies,^24^ are particularly concerning in the context of *A. baumannii*. The phenotype-agnostic approach to sampling in this study allowed us to determine that the majority (80·8%) of *A. baumannii* in this ICU were CRAB. We observed an increase in the magnitude of carbapenem resistance in this ICU, with MIC_50_/MIC_90_ values doubling between our 2019 and 2021 studies. The increase in carbapenem resistance levels was driven by the emergence of ST164, which co-produced NDM-1 and OXA-23 carbapenemases. Despite the increase in carbapenem resistance, all isolates obtained in 2021 remained sensitive to colistin and tigecycline, though monitoring CRAB populations for resistance to these last-resort antibiotics should remain a high priority.

Following IPC interventions, we observed a decrease in the numbers of CRAB isolated from patients and from the ICU environment, and saw a reduction in both the total number of clinical isolates (from 19 in 2019 to 13 in 2021) and the number of patients from which clinical isolates were obtained: 12/140 patients (8·6%) in 2019, and 10/131 patients (7·6%) in 2021. However, IPC practices remained imperfect, with multiple examples of GC2 CRAB clusters being imported to the ICU before spreading to different bed units or rooms and being acquired by patients, highlighting the significant challenges associated with prevention of CRAB transmission in hospital wards once it has colonised the hospital environment. By using a highly-stringent 0-cgSNP cut-off when assessing putative transmission events over the course of this study, we have unambiguously implicated environmental persistence in the acquisition of CRAB by ICU patients and ultimately the development of nosocomial infections. New data presented here adds to our previous findings that suggested GC2 clusters were being imported to the ICU regularly from external sources. We hypothesise that other wards within this hospital, as well as patients transferred from other hospitals, serve as a source for incoming CRAB strains. If this is the case, hospital-wide IPC measures or highly stringent screening and isolation of CRAB-positive patients prior to ICU admission will be required to ultimately address CRAB imports.

A striking finding here was the reduction in prevalence of GC2 amongst CRAB isolates, falling from 99·5% in 2019 to 50·8% in 2021. The remainder of the 2021 CRAB population was comprised of a highly-clonal set of ST164 isolates that appear to have been diversifying *in situ* since mid-2020. This study has therefore added further context to the recent recognition of the emergence of ST164 as an international *A. baumannii* clone,^20^ with our data specifically highlighting this clone’s capacity to establish itself and persist in an ICU environment, even in direct competition with clusters of the highly successful GC2. While ST164 caused fewer infections than GC2 over this study period, its presence in one clinical specimen, and in clinical specimens collected previously in this ICU and elsewhere,^19,20^ along with its high carbapenem resistance levels, suggest that it is a lineage that requires surveillance and further characterisation.

The increase in CRAB diversity between 2019 and 2021, including the appearance of ST164 and the diversification of GC2, was likely a product of regular imports to the ICU over an approximately 20-month period. However, it would be interesting to determine whether IPC interventions have introduced new selective pressures and contributed to population turbulence. If so, these environmental conditions might select for fitter lineages that can better establish persistence in hospital environments. Environmental persistence might also be associated with a notable trade-off in virulence, as seen by the skew in isolates causing clinical infections here. Under such conditions, IPC measures might have differential effects at the strain level, and effective responses will require informed, dynamic IPC strategies. Experimental characterisation of diverse CRAB lineages will be needed to assess their variable ability to persist in hospital environments and ultimately infect patients. The GC2 cluster C25, which was persistent in this ICU and prominent amongst clinical isolates over the course of this study (10/13; 76·9%), is an intriguing target for future laboratory studies that might address these questions.

## Data Availability

All sequence data produced in this study are available online via NCBI under accession number PRJNA1034534

## Acknowledgements

This work was undertaken as part of the DETECTIVE research project funded by the Medical Research Council (MR/S013660/1), and the National Natural Science Foundation of China (U22A20338, 32270183, 81861138054, 32011530116). W.v.S was also supported by a Wolfson Research Merit Award (WM160092). AM is also supported by the National Institute for Health and Care Research (NIHR) Birmingham Biomedical Research Centre (NIHR203326).

