## Supplementary Figures for "Changes to an intensive care unit *Acinetobacter baumannii* population following COVID-19 disruptions and targeted infection prevention interventions"

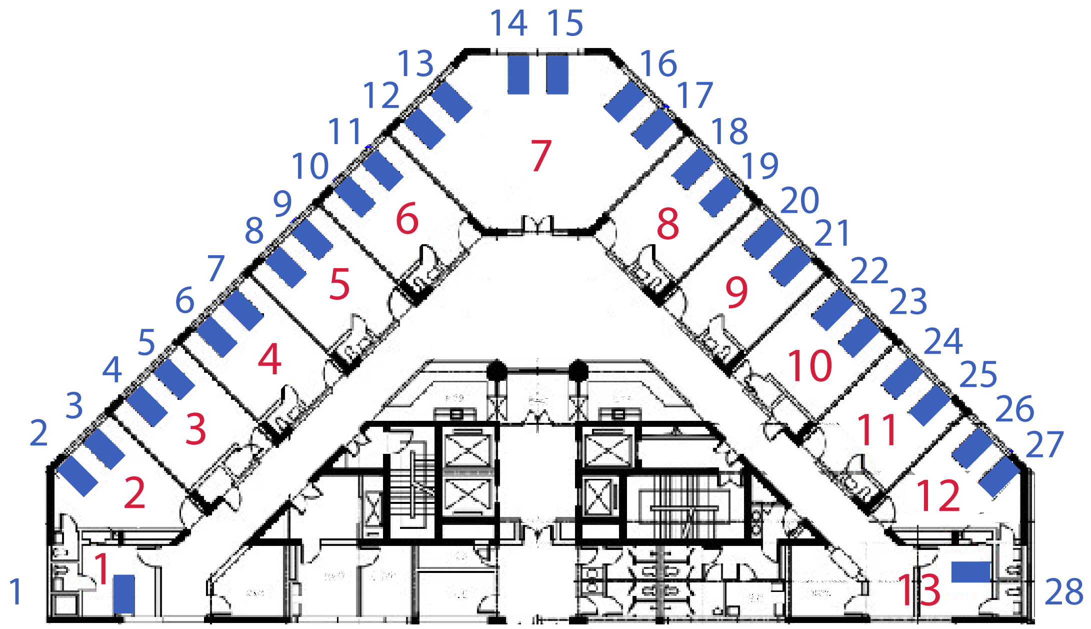


**Figure S1:** Intensive care unit (ICU) map. Floorplan of the ICU where this study was conducted. Blue numbers refer to bed units and red numbers to rooms.

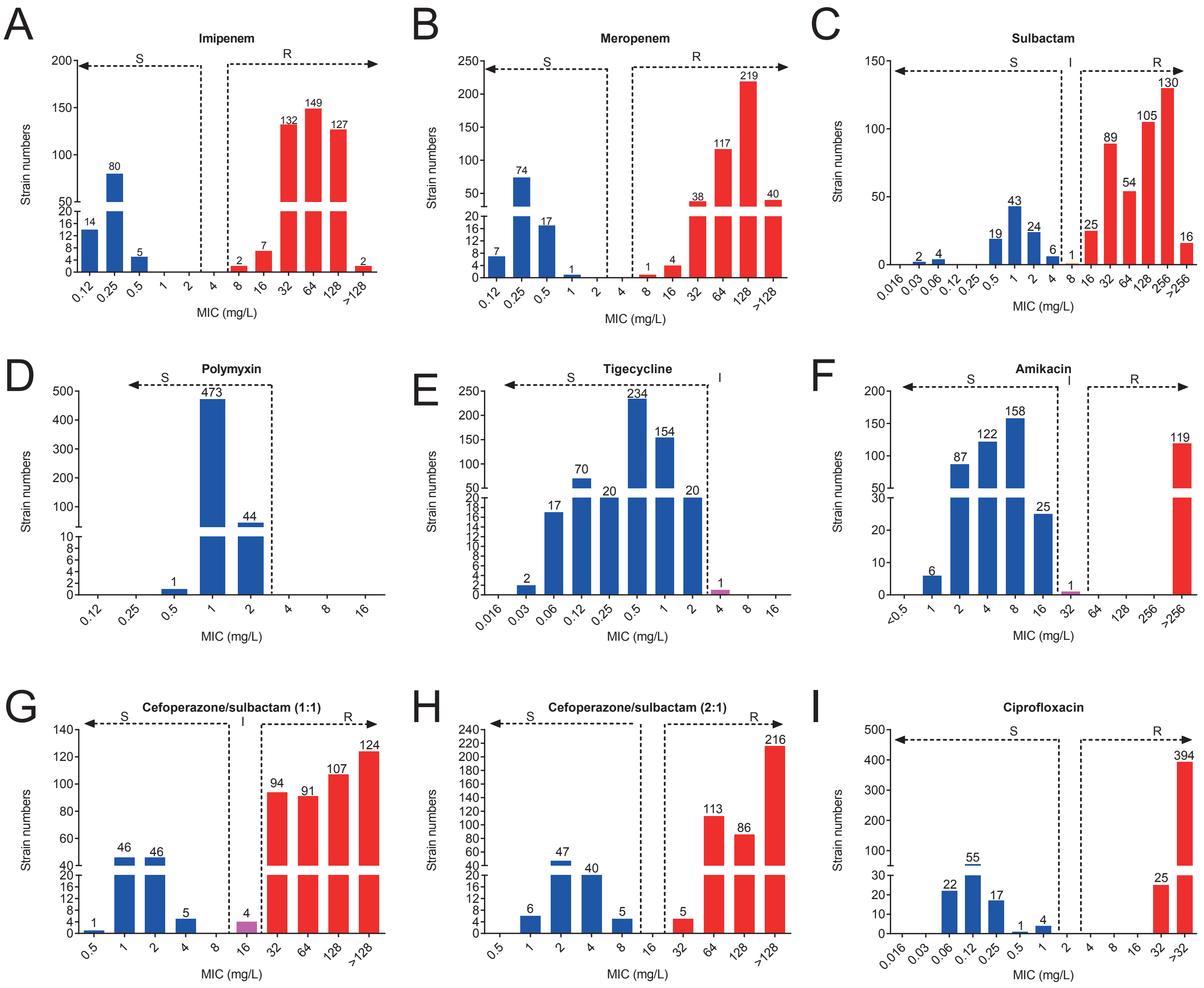


**Figure S2:** Distribution of MICs for 9 antimicrobial agents against of 518 *A. baumanii*. A) imipenem; B) meropenem; C) sulbactam; D) polymixin; E) tigecycline; F) amikacin; G) cefoperazone/sulbactam (1:1); H) cefoperazone/sulbactam (2:1); I) ciprofloxacin.

Notes: 1) As there is no tigecycline breakpoint for *Acinetobacter*, MICs were interpreted following the FDA guidelines for Enterobacterales (with MICs ≤ 2 mg/L denoting susceptibility and ≥ 8 mg/L denoting resistance). 2) As there is no breakpoint for cefoperazone/sulbactam, results were interpreted according to the CLSI 2021 ampicillin/sulbacta guidelines.

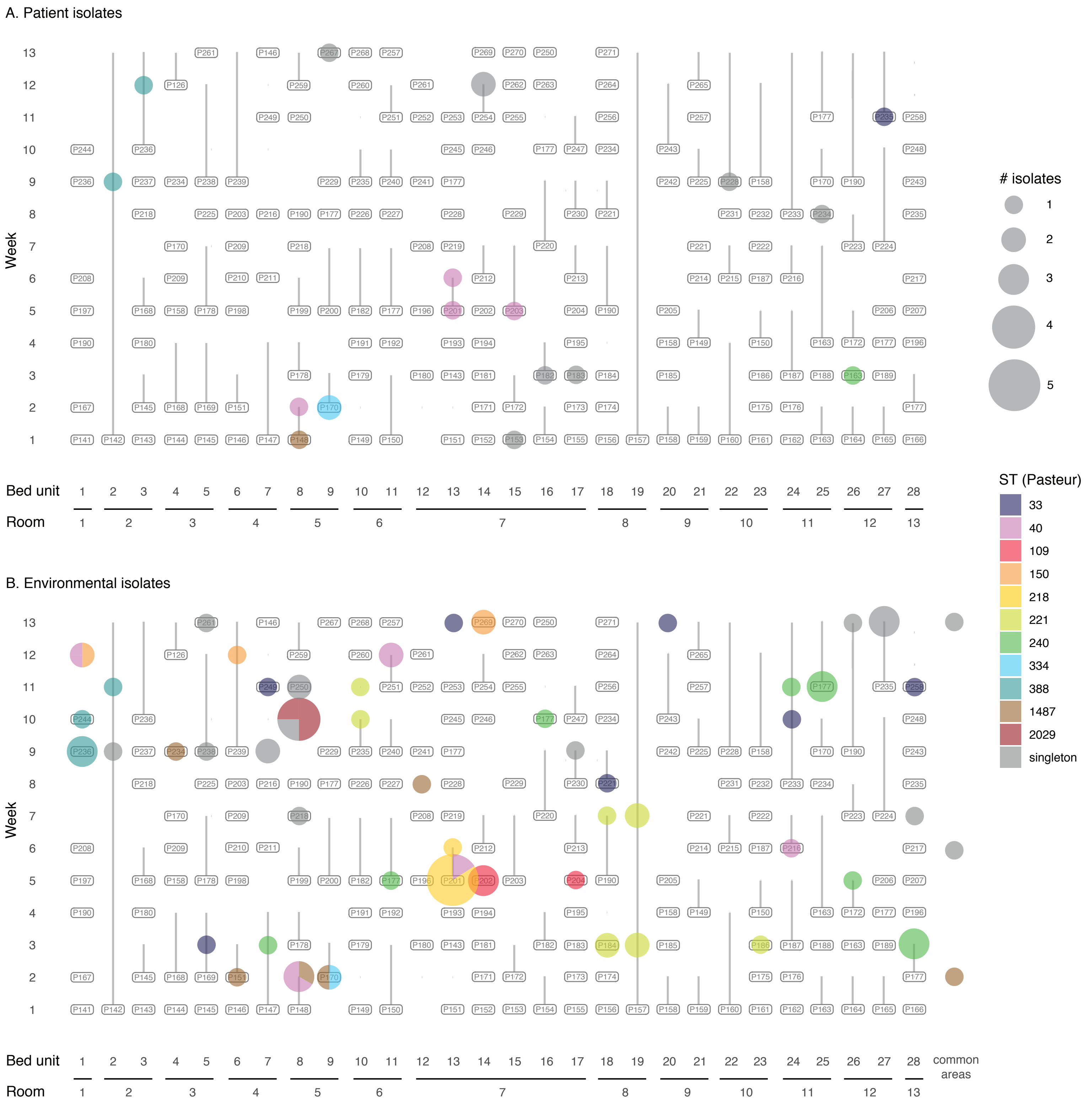


**Figure S3:** Spatiotemporal distribution of carbapenem-sensitive *Acinetobacter baumannii* (CSAB) in A) patient and B) environmental samples. Coloured circles correspond to the sequence type (ST) and number of isolates collected from each bed unit or patient at each time point, as outlined in the key to the right of the figure.

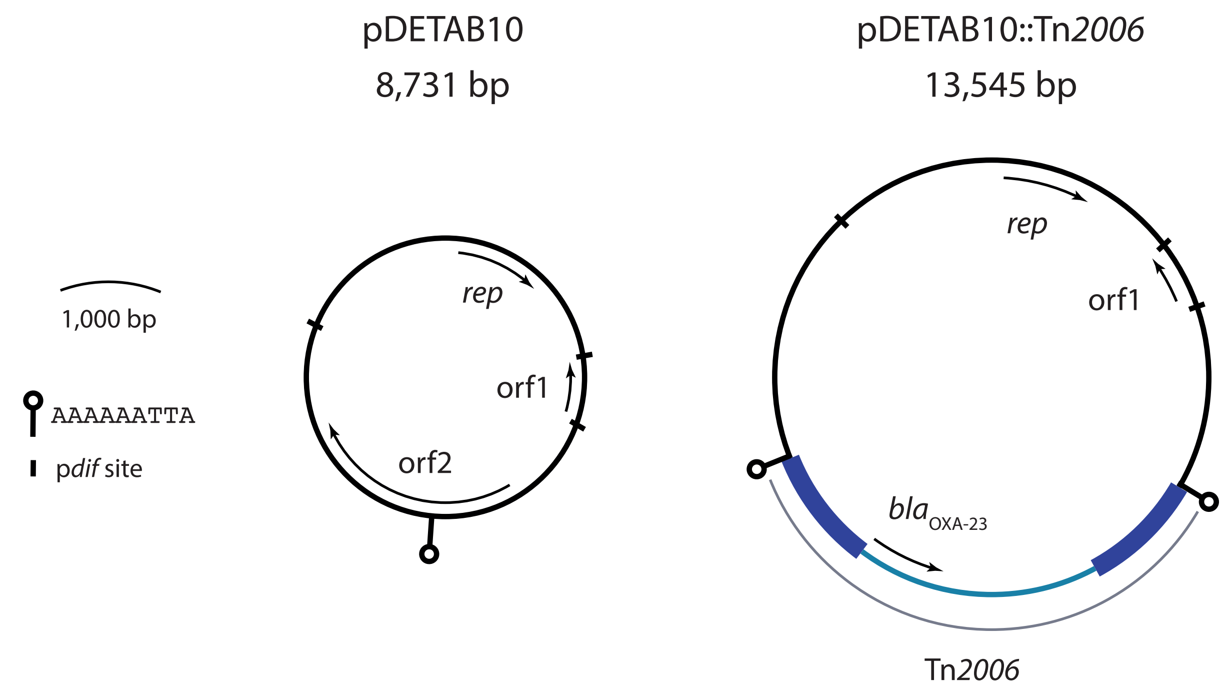


**Figure S4:** Acquisition of Tn*2006* by pDETAB10. Scaled schematic diagrams of the R3-T1 plasmids pDETAB10 and pDETAB10::Tn*2006*. The plasmid backbone is shown as a black line, the insertion sequence IS*Aba1* as navy boxes, and the Tn*2006* passenger segment as a light blue line. Open reading frames are shown as labelled arrows. The locations of target site duplications and p*dif* sites are indicated as shown in the key to the left of the figure.
