## Supplementary material for "Changes to an intensive care unit *Acinetobacter baumannii* population following COVID-19 disruptions and targeted infection prevention interventions": Table S1

**Table S1:** IPC interventions

| **Focus** | **Intervention(s)** |
| --- | --- |
| Patients | ✓“Water-free” patient care:   1. Patients who will stay in ICU the for only 1 day or only for postoperative monitoring: use ordinary wet wipes for care; 2. Patients who will stay in the ICU for >1 day but have no evidence of multi-drug resistant bacterial infection: use dry cleaning liquid; 3. Patients who have multi-drug resistant bacterial infections: use dry cleaning liquid + chlorhexidine wipes;   ✓Active surveillance of patients for CRAB on admission and every other week: positive patients would be sent to isolation room.  ✓Install two disinfection devices for patients’ urine and liquid waste. |
| ICU environment and equipment | ✓Increase the frequency of equipment surface cleaning in every ward: one towel per bed unit.  ✓Replace the keyboard cover every day and send it to the supply room for cleaning and disinfection.  ✓Clean all domains and equipment of the ward when the patients discharge.  ✓Increase cleaning of crash trolleys and computers in nurse station. |
| ICU sinks | ✓Sinks in patient rooms no longer in use, but have not been removed. |
| Hospital staff | ✓Enhanced medical staff education on contact precautions.  ✓Hand hygiene enforcement and compliance monitoring: no-touch quick-drying hand disinfectant placed in every room, proper use of gloves and isolation gowns.  ✓Build a hand hygiene compliance management system: nurses wear a sensor, nurses will go for handwashing when the system detect no their handwashing record. |
