## Supplementary material for "Changes to an intensive care unit *Acinetobacter baumannii* population following COVID-19 disruptions and targeted infection prevention interventions": Table S2

**Table S2:** Clinical data of patients that were in the ICU during this study.

| **Item** | **Number of patients** | **percentage** |
| --- | --- | --- |
| **Sex** |  |  |
| Male | 79 | 60.3% (79/131) |
| Female | 52 | 39.7% (52/131) |
| **Age (Years)** |  |  |
| 0-5 | 0 | 0 (0/131) |
| 6-64 | 53 | 40.5% (53/131) |
| ≥65 | 78 | 59.5% (78/131) |
| **Patients’ ward source** |  |  |
| Admission ICU directly | 91 | 62.3% (91/146^a^) |
| General Surgery | 26 | 17.8% (26/146) |
| Infectious Disease | 6 | 4.1% (6/146) |
| Orthopedics | 4 | 2.7% (4/146) |
| Gastroenterology | 3 | 2.1% (3/146) |
| Neurology | 3 | 2.1% (3/146) |
| Respiratory Medicine | 3 | 2.1% (3/146) |
| Hematology | 3 | 2.1% (3/146) |
| Others | 7 | 4.8% (7/146) |
| **Length of ICU stay** |  |  |
| 0-10 days | 78 | 53.4% (78/146) |
| 11-20 days | 31 | 21.2% (31/146) |
| 21-30 days | 12 | 8.2% (12/146) |
| > 31 days | 25 | 17.1% (25/146) |
| **Antimicrobials used in the ICU** |  |  |
| Carbapenems^b^ | 25 | 19.2% (25/131) |
| Cefoperazone/Sulbactam | 19 | 14.5% (19/131) |
| Piperacillin/Tazobactam | 53 | 40.5% (53/131) |
| Levofloxacin | 6 | 4.6% (6/131) |
| Moxifloxacin | 2 | 1.5% (2/131) |
| Polymyxin B | 6 | 4.6% (6/131) |
| Tigecycline | 8 | 6.1% (8/131) |
| Unused | 7 | 5.3% (7/131) |

^a^146: Eight patients admitted into the ICU and discharged from the ICU multiple times during the three-month sampling period. Thus, the number is 146 and not 131.

^b^Imipenem, memropenem, ertapenem.
